# Psychiatric, cognitive, psychosocial, and neurological outcomes of chimeric antigen receptor T-cell therapy: protocol for a prospective study

**DOI:** 10.1101/2023.09.03.23294983

**Authors:** Valeriya Kuznetsova, Harsh Oza, Hannah Rosenfeld, Carmela Sales, Samantha van der Linde, Izanne Roos, Stefanie Roberts, Fiore D’Aprano, Samantha M Loi, Mark Dowling, Michael Dickinson, Tomas Kalincik, Simon J Harrison, Mary Ann Anderson, Charles B Malpas

**Author notes:** Corresponding author: Valeriya Kuznetsova, Contact address: Clinical Outcomes Research Unit, Level 4 East, Royal Melbourne Hospital, 300 Grattan Street, Parkville VIC 3052, E. Equally credited authors (Mary Ann Anderson, Charles B Malpas).

## Abstract

**Background:** Immune effector cell-associated neurotoxicity syndrome (ICANS) is a common side-effect of chimeric antigen receptor T-cell (CAR-T) therapy, with symptoms ranging from mild to occasionally life-threatening. The psychiatric, cognitive, psychosocial, and neurological sequalae of ICANS are diverse and not well-specified, posing a challenge for diagnosis and management. The recovery trajectory of the syndrome is uncertain. Psychiatric, cognitive, psychosocial, and neurological status is rarely examined in this population pre-therapy, adding a layer of complexity to specifying symptoms pertinent solely to CAR-T treatment.

**Aims:** The aim is to investigate psychiatric, cognitive, psychosocial, and neurological outcomes in patients after CAR-T therapy, particularly among those who develop ICANS. The project will establish a comprehensive pre-treatment baseline and will longitudinally monitor for therapy-associated change.

**Methods:** A prospective longitudinal study of all adult patients in a single Australian haematology service undergoing CAR-T therapy. Neuropsychological and neurological examinations occur prior to CAR-T, and patients are reviewed during the acute post-treatment period, 28 days, 6 months, and 12 months post-infusion. Data will be sourced from objective psychometric measures, clinical examinations, self-report questionnaires, and accounts of subjective cognitive complaint to capture a broad spectrum of dysfunction and its daily functional impact.

**Conclusions:** We present a protocol of a research study that will describe the neurocognitive features specific to ICANS, characterise the underlying syndrome, identify predictors of differential post-infusion outcomes, and contribute to optimising the overall management of CAR-T patients. The protocol will serve as the basis of guidance regarding clinical and paraclinical follow-up of patients undergoing CAR-T cell therapy.

## Introduction

### Background

Chimeric antigen receptor T-cell (CAR-T) therapy is an immunotherapy that modifies T-cells to target and eliminate cancer cells and is fast becoming a standard of care for some relapsed or refractory haematological malignancies.(1,2) Cytokine release syndrome (CRS) and immune effector cell-associated neurotoxicity syndrome (ICANS) are common side-effects of CAR-T.(2,3) ICANS has been described in as high as over 60% of patients depending on a CAR-T product. The most commonly reported consequence is encephalopathy, which can range from mild cognitive and neurological deficits to obtundation, stupor, and even coma.(1,4) The onset tends to vary from days to weeks post-infusion, with limited research available on neurocognitive recovery trajectory.(5) Although the literature has documented a diverse range of clinical features of ICANS, the underlying syndrome has not been well-characterised.(6)

This paper describes a protocol of an Australian prospective longitudinal study of adult patients in the haematology service undergoing CAR-T therapy. The aim is to investigate psychiatric, cognitive, psychosocial, and neurological outcomes of the treatment, with an emphasis on characterisation of acute neurocognitive features of ICANS. The study will determine a set of cognitive instruments necessary to identify and monitor the syndrome acutely and investigate predictors of differential outcomes post-infusion. Recovery will be examined from an acute, sub-acute, and long-term perspective. This paper outlines the rationale, design, and methodology of the research protocol.

### Clinical features of ICANS

Accurate recognition of ICANS is essential for timely intervention and adequate medical management. Description of clinical presentation varies across studies, and can include focal weakness, apraxia, tremor, aphasia, perseverative and hesitant speech, dysgraphia, disorientation, impaired attention, memory dysfunction, seizures, hallucinations, behaviour disturbance, and emotional lability.(3,5–8) There has been, however, little consensus regarding a syndrome underpinning these symptoms, which poses a challenge for early detection, classification of severity, and recovery monitoring. Neurotoxicity is typically treated with corticosteroids, which have the drawback of potentially reducing efficacy of CAR-T cells.(5) Understanding the syndrome evolution could help optimise dosage and duration of corticosteroid administration, subsequently improving therapy outcomes.

The commonly adopted measure of ICANS is the immune effector cell-associated encephalopathy (ICE) score from the American Society for Transplantation and Cellular Therapy.(9) The ICE is a 10-point instrument comprising brief questions on orientation, attention, handwriting, simple naming, and ability to follow simple commands. Severity classification incorporates the ICE score with an assessment of level of consciousness, motor function, evidence of seizures and elevated intracranial pressure.(1,5,9) Although the scale is easy to administer at regular intervals, it might not be sensitive to more subtle dysfunction. For example, Herr et al.(10) described a series of patients in whom subtle neurocognitive symptoms predated decrease in the ICE score. Thus, the ICE might not adequately recognise early signs of mild impairment or capture recovery to complete resolution. Understanding syndromology of neurotoxicity is the first step in development of new diagnostic instruments and evidence-based management strategies.

### Baseline significance

One of the key challenges in evaluating neurocognitive status post-CAR-T is distinguishing new symptoms from pre-existing dysfunction. Both disease itself and cancer treatments are commonly associated with neurological and cognitive presentation. Patients become eligible for CAR-T if their cancer is refractory to or has progressed following prior lines of systemic therapy or stem cell transplantation. Many have been exposed to central nervous system (CNS) penetrating agents. Schroyen et al.’s(11) systematic review demonstrated high prevalence of chemotherapy-induced leukoencephalopathy in cancer patients, persisting for years following active treatment. A colloquial term ‘chemofog’ describes transient cognitive dysfunction post-chemotherapy, which can affect processing speed, attention, and secondarily impact learning, memory, and executive function.(12) Subjective cognitive complaints are prevalent in the oncology population, with physical and psychological factors (e.g., poor sleep, fatigue, pain, anxiety, and depression) commonly contributing to disruption in cognition.(13–15) Thus, to adequately characterise ICANS it is necessary to evaluate baseline cognitive, psychological, and neurological status in patients pre-CAR-T.

### Recovery trajectory

Although ICANS is understood as an acute event typically resolving within several weeks, persisting symptoms have been reported. Research examining neurocognitive recovery has been limited.(5,16) Maillet et al.(17) prospectively examined neurocognitive outcomes of neurotoxicity 6-12 months after CAR-T in 27 adult patients with relapsed B-cell lymphoma. Prior to the infusion, participants underwent neurological and neuropsychological examinations and completed self-report questionnaires on mental health and subjective cognitive complaint. Forty-four percent experienced acute neurotoxicity, and those who survived without disease progression were reviewed 5-12 months post-treatment. There was no difference in participants’ neurocognitive status at review compared to baseline, suggesting that ICANS had resolved. The authors also discovered a reduction in reports of anxiety and subjective cognitive concerns. While these findings are consistent with the hypothesised acute nature of neurotoxicity, a large proportion of the cohort was excluded from follow-up due to disease progression. It is not clear if these patients followed the same trajectory. A wide follow-up interval also limits conclusions that can be drawn regarding specifics of ICANS duration.

Wang et al.(18) conducted a cross-sectional observational study of 60 patients in the first year post-CAR-T (28 were <30 days, 13 were 30-90 days, and 19 were >90 days) to capture patient-reported concerns. Among others, commonly reported symptoms included pain, fatigue, sleep disturbance, drowsiness, lack of energy, malaise, weakness, headache, and difficulty concentrating. In patients who were >30 days post-infusion, grade 2-4 CRS and/or ICANS was associated with greater reports of physical symptoms, sadness, irritability, difficulty speaking, and interference with enjoyment of life compared to patients who experienced no or low-grade side-effects. The authors did not exclude patients with disease progression from their analyses and did not have information on symptom profiles prior to treatment. Thus, distinguishing CAR-T related contribution from possible effects of disease and other treatment regiments is difficult. These findings provide valuable insight into subjective experience of CAR-T patients and their quality of life. Self-report questionnaires could be used as screening instruments for patients who could benefit from clinical consultation and subsequent intervention.

### Risk of neurotoxicity

Identifying patients at risk of neurotoxicity is paramount for adequate medical management pre- and post-CAR-T. Established risk factors for developing ICANS include older age, active CNS disease, CAR-T cell dose and product (e.g., higher incidence has been reported following axicabtagene ciloleucel), early onset of CRS, and high disease burden.(16,19) The literature has suggested that serum markers, such as procalcitonin, ferritin, IL-6, C-reactive protein, and lactate dehydrogenase might also predict development of ICANS, however, the evidence is mixed.(19–21) There are no known cognitive markers predictive of development or severity of neurotoxicity. Identifying new risk factors and prodrome markers of ICANS would help establish uniform guidelines for medical care and implementing preventative strategies for those at high risk.(7)

## Project Aims

The proposed research aims to investigate neurocognitive, psychiatric, and psychosocial outcomes of CAR-T therapy. By establishing a comprehensive neuropsychological and neurological baseline pre-treatment, we will define acute neurocognitive features specific to ICANS and severe CRS. Characterization of the baseline cognitive status will extend beyond psychometric investigation to evaluating semiology of the cognitive complaint. Recordings of neuropsychology interviews will allow for a comprehensive analysis of subjective cognitive concerns that might negatively impact patients’ daily life. The project will examine objective neurocognitive changes and early signs associated with neurotoxicity, as well as define a set of instruments necessary to detect and monitor ICANS. Neuropsychological examinations will be completed by a clinical neuropsychologist, who will distinguish primary cognitive impairment underpinned by CAR-T related processes from secondary cognitive dysfunction resulting from psychopathology, poor sleep, fatigue, or pain. The study will also investigate predictive factors for developing neurotoxicity to facilitate close monitoring and preventative management of patients who are at high risk. Finally, both acute and long-term recovery will be examined post-CAR-T from a neurological, cognitive, and psychosocial perspective. It is hoped that the research will contribute to optimising the overall management of CAR-T patients and improve recognition and referral pathways for neurocognitive dysfunction in this cohort.

## Method

### Design

The study is a prospective longitudinal cohort study of adult patients undergoing CAR-T therapy at Peter MacCallum Cancer Centre (PMCC) haematology service. Patients are examined at baseline, prior to CAR-T, and are reviewed during the acute post-treatment period, as well as 28 days, 6 months, and 12 months post-infusion. The project also includes cross-sectional recruitment of a matched healthy comparator group. The authors assert that all procedures contributing to this work comply with the ethical standards of the relevant national and institutional committees on human experimentation and with the Helsinki Declaration of 1975, as revised in 2008. All procedures involving human subjects/patients were approved by the Peter MacCallum Cancer Centre Human Research Ethics Committee (21/145).

### Participants

#### Clinical cohort

The study will include patients planned for CAR-T therapy if they are at least 18 years of age and do not have an intellectual disability or a history of significant neurological illness. All patients complete a routine specialist cognitive assessment by a clinical neuropsychologist and a neurological examination by a treating neurologist pre- and post-CAR-T. Participants are recruited via the Centre of Excellence for Cellular Immunotherapy at PMCC. The findings from neuropsychological assessments are included only for those proficient in English. Data from baseline examinations is included for patients who are planned for commercial CAR-T products or for clinical trials, however, post-infusion follow-up sample will only comprise those who receive commercial products. Participants provide written informed consent. A waiver of the requirement for consent was approved by the Ethics Committee to access routine clinical data for patients who received CAR-T prior to commencement of this study (between May 2021 and January 2023).

#### Healthy controls

To collect optimal demographically adjusted psychometric normative data, healthy participants are recruited through hospital-based advertising to complete a single specialist cognitive assessment. Eligible participants are over 18 years of age, are proficient in English, have adequate uncorrected or corrected eyesight to perceive examination material, do not have past or present cancer history, significant neurological or neurodevelopmental history, and do not have a current significant psychiatric condition. Healthy controls are prospectively consented and are matched to the clinical cohort on age, sex, and education.

### Power and sample size

Based on clinical experience in the CAR-T program, the following project aims to recruit 100 CAR-T patients. Proportion of patients who develop ICANS following CAR-T therapy has been reported to range from 23 to 67 percent in lymphoma cases and 40 to 62 percent in patients with leukaemia.(1,8) Taking a conservative approach, we estimate that approximately 35 percent of our patients will develop ICANS of any grade. Thus, we expect to have 35 patients in the ICANS group and 65 patients in the non-ICANS group. This represents the expected number of patients to pass through the CAR-T program as part of routine clinical care. Based on this samples size, the majority of analyses will be sensitive to small effect sizes. For example:

- For bivariate Pearson’s correlations (e.g., for examination of associations between continuous variables at baseline), this sample size would render the analyses sensitive to effect sizes of *r* = 0.28 (power = 80%, alpha = 5%, sample size = 100, two tailed).
- For dependent samples t-tests (e.g., for examination of the mean change from baseline to post-CAR-T), this sample size would render the analyses sensitive to small effect sizes of *d* = 0.28 (power = 80%, alpha = 5%, sample size = 100, two tailed).
- For independent samples t-tests (e.g., for comparisons of means between the anticipated 35 ICANS patients and the 65 non-ICANS patients) this sample size would render the analyses sensitive to medium effect sizes of *d* = 0.59 (power = 80%, alpha = 5%, sample size = 100, two tailed).
- For analysis of covariance (ANCOVA) for comparison across neuropsychological diagnostic groups with age included as a covariate, the sample size would render the analysis sensitive to a small effect size of *f* = 0.09 (power = 80%, alpha = 5%, sample size = 100, two tailed).

### Procedure

A standard care neuropsychological assessment takes approximately 2 hours and includes (1) self-report online questionnaires on health, psychological wellbeing, cognitive dysfunction, social participation, and quality of life, (2) clinical interview to elicit a cognitive complaint (if any), and (3) psychometric assessment of cognition. Prospectively consented participants are asked permission to audio record the clinical interview for subsequent transcription. A standard care neurological assessment includes (1) clinical interview to collect medical history and elicit neurological complaint, (2) physical neurological examination, and (4) a screening measure of objective cognitive impairment. Table 1 summarises the schedule of routine assessments for the clinical cohort, including self-report questionnaires and psychometric instruments. Modified ad hoc inpatient examinations are completed during acute post-infusion period, as clinically indicated. In line with standard of care, neurotoxicity is monitored by inpatient ward staff via the ICE score.(9) Patients also undergo neuroimaging investigations as clinically indicated.

**Table 1.**
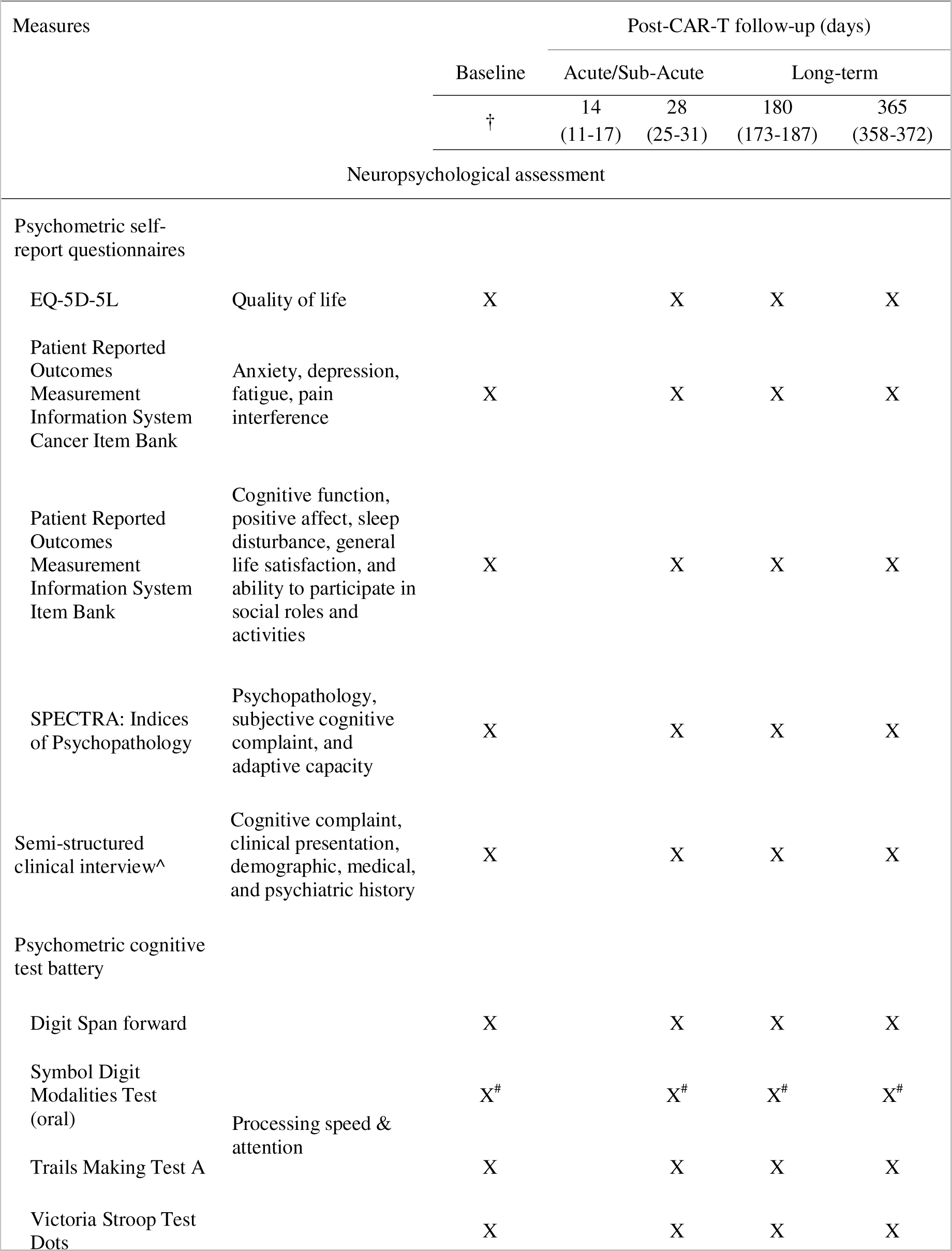
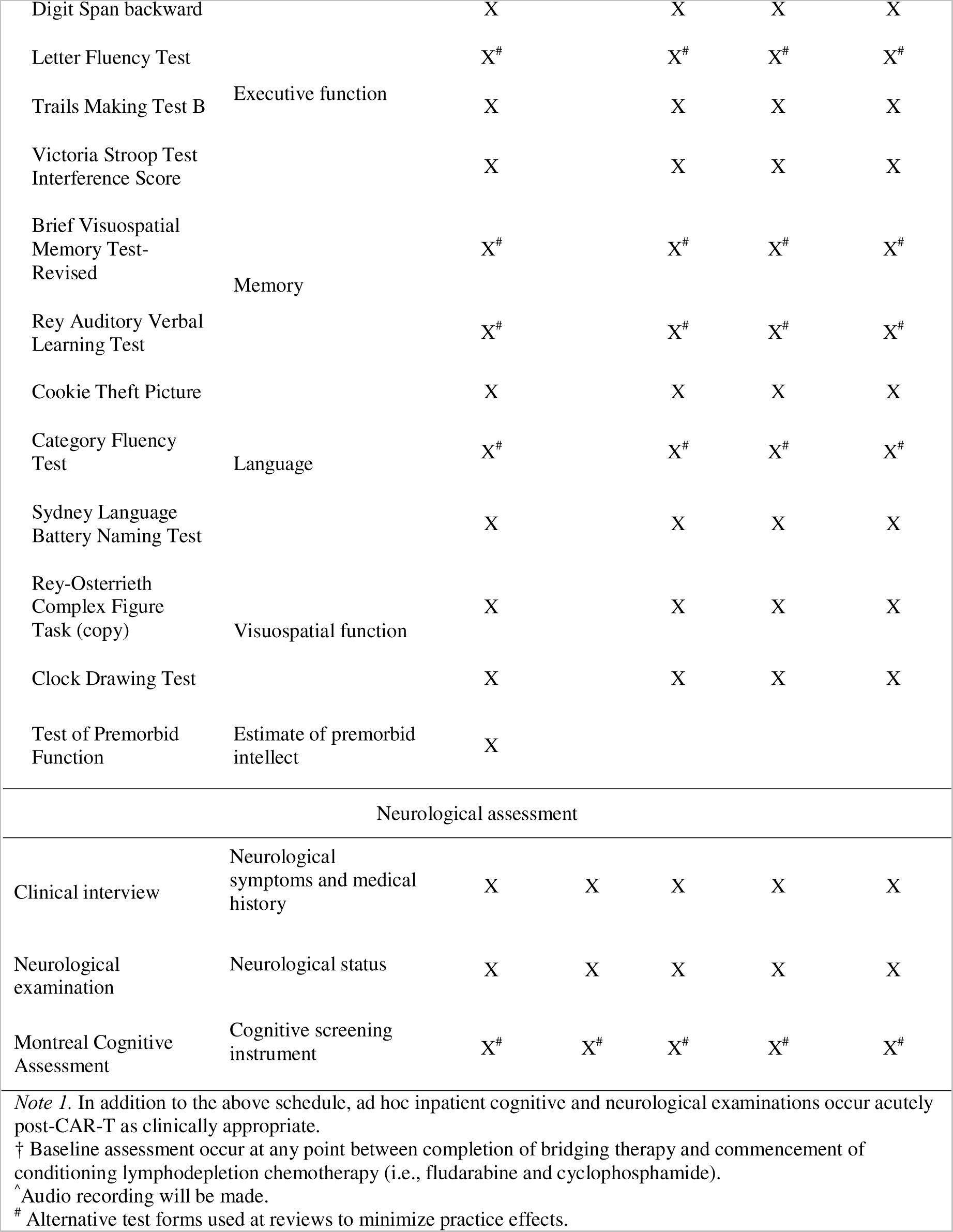
Schedule of routine assessments.

Medical history and information on demographic, disease, and treatment characteristics is obtained from patient charts and clinical interviews. Neuropsychology and neurology examinations are primarily conducted at PMCC and the Royal Melbourne Hospital (RMH), Department of Neurology, with some follow-ups conducted via telehealth to minimise travel burden on participants. Healthy participants complete a single neuropsychological assessment either in person or via telehealth.

### Measures

#### Questionnaires

Psychometric self-report questionnaires encompass domains of psychological, physical, and psychosocial function relevant to our patients. Information collected via self-report measures contributes to a holistic understanding of CAR-T patients’ experience and aids a clinical distinction between primary and secondary cognitive dysfunction.

The SPECTRA Indices of Psychopathology(22) measures psychopathology, subjective cognitive complaint, and adaptive capacity. The instrument comprises 96 statements, to which participants respond on a 5-point Likert scale (1 = not at all true; 5 = completely true). The SPECTRA has demonstrated good internal consistency in psychiatric patients (*a* = 0.74-0.95) and correlates highly with other measures of psychopathology.(22,23)

Instruments from the cancer bank of the Patient Reported Outcomes Measurement Information System (PROMIS)(24) are included to examine aspects of psychosocial functioning and physical symptoms specific to the oncology population. These were developed based on interviews with cancer patients. From the battery, we selected measures of anxiety, depression, fatigue, and pain interference. We also included measures from the broader PROMIS repository on subjective cognitive function, positive affect, sleep disturbance, general life satisfaction, and ability to participate in social roles and activities. All questionnaires are completed online as computerized adaptive test (CAT). Participants respond to items on a Likert scale, and the number of questions varies depending on response patterns.

To examine the overall quality of life, participants complete the brief EQ-5D-5L questionnaire developed by the EuroQol Group.(25) The instrument examines health status across five dimensions: mobility, self-care, usual activities, pain/discomfort, and anxiety/depression. For each item, participants select out of 5 statements (1 = no problems; 5 = unable to/extreme problems). Overall health is rated on the visual analogue scale 1-100.

#### Semi-structured clinical interview

All participants complete a clinical interview by a clinical neuropsychologist to elicit a subjective cognitive complaint, examine clinical presentation, and collect demographic and medical history. Prospectively consented participants are asked permission to audio record baseline interviews to enable a focus on the descriptive quality of speech to characterise common cognitive complaints in this cohort. Clinicodescriptive analysis of subjective cognitive complaint will contribute to describing baseline neuropsychological status specific to haematology patients compared to the healthy controls. Components of psychometric examination and self-reported information will encompass quantitative analyses to clarify the mechanisms underlying typologies of cognitive complaint. This semi-structured interview process was developed using clinical experience to mirror the standard care interview, and in alignment with previous interviews constructed to elicit detailed cognitive complaint.(26) The interview is structured to probe circumstances in which cognitive dysfunction is likely to occur while obtaining features of frequency, contextualization, and recovery from cognitive failure. Recorded clinical interviews will be transcribed and analysed.

#### Psychometric cognitive examination

A battery of psychometric cognitive instruments was selected to capture a wide range of cognitive dysfunction possible in this cohort. The psychometric battery examines five cognitive domains in accordance with the recommendations from the International Cognition and Cancer Task Force (ICCTF).(27) The processing speed and attention domain is measured with the Trail Making Test Part A,(28,29) Victoria Stroop Test Dots,(30) Symbol Digit Modalities Test,(31) and Digit Span forward from the Wechsler Adult Intelligence Scale 4th edition (WAIS-IV).(32) The executive function domain comprised the Trail Making Test Part B,(28,29) Letter Fluency Test from the Delis–Kaplan Executive Function System (D-KEFS),(33) Victoria Stroop Test interference score,(30) and Digit Span backward from the WAIS-IV.(32) The memory domain was measured with the Rey Auditory Verbal Learning Test(34) and the Brief Visuospatial Memory Test-Revised.(35) The language domain included the Category Fluency Test from the D-KEFS(33) and the Sydney Language Battery Naming Test.(36) The visuospatial function domain was measured using the Rey-Osterrieth Complex Figure Test Copy.(37) Participants also complete the Clock Drawing Test,(38) which has complex visuospatial demands. The task, however, is also sensitive to impairments in other areas, such as semantic and executive dysfunction. The Test of Premorbid Function(39) is administered to estimate premorbid intellect. Table 2 outlines details of all cognitive instruments.

**Table 2.**
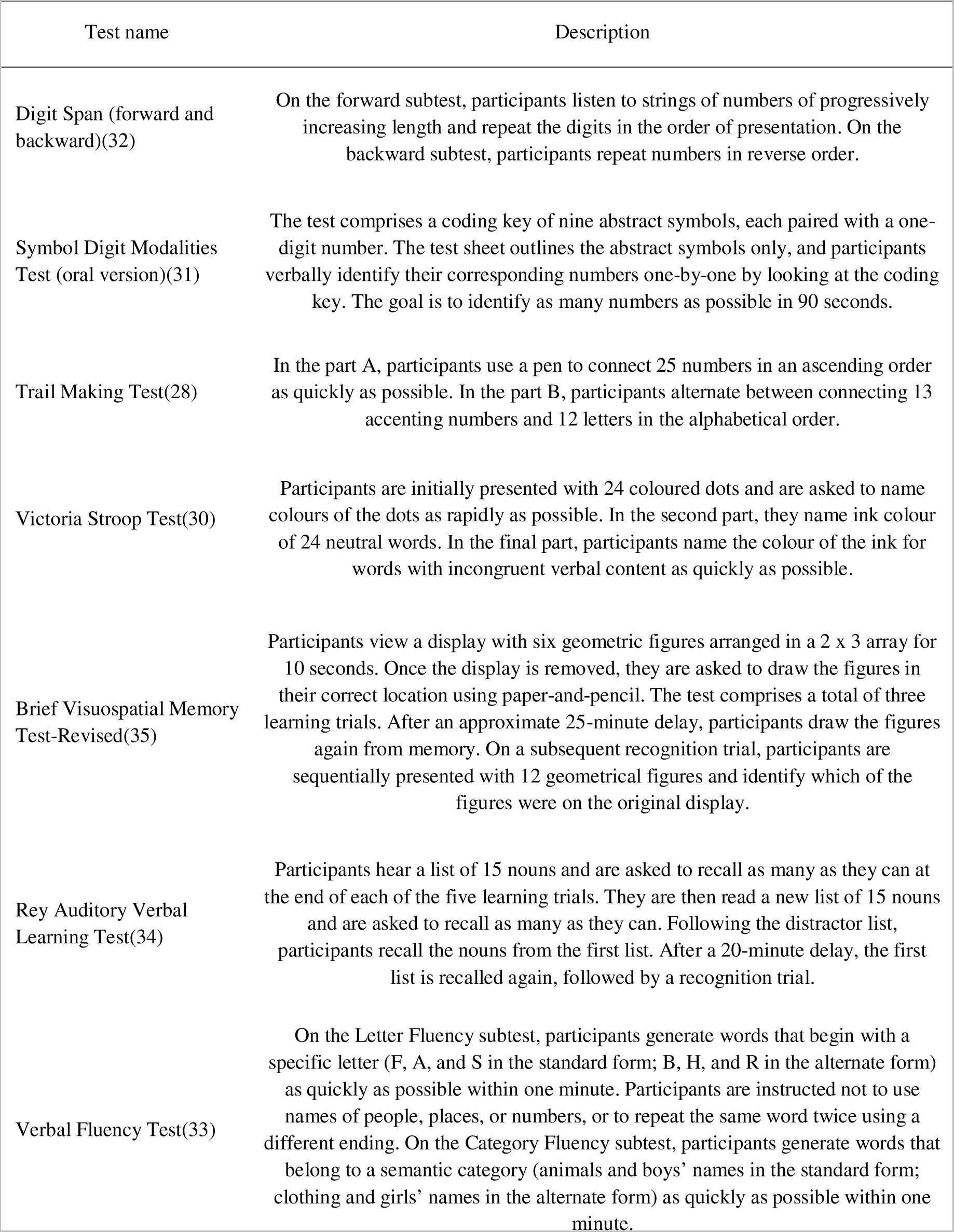
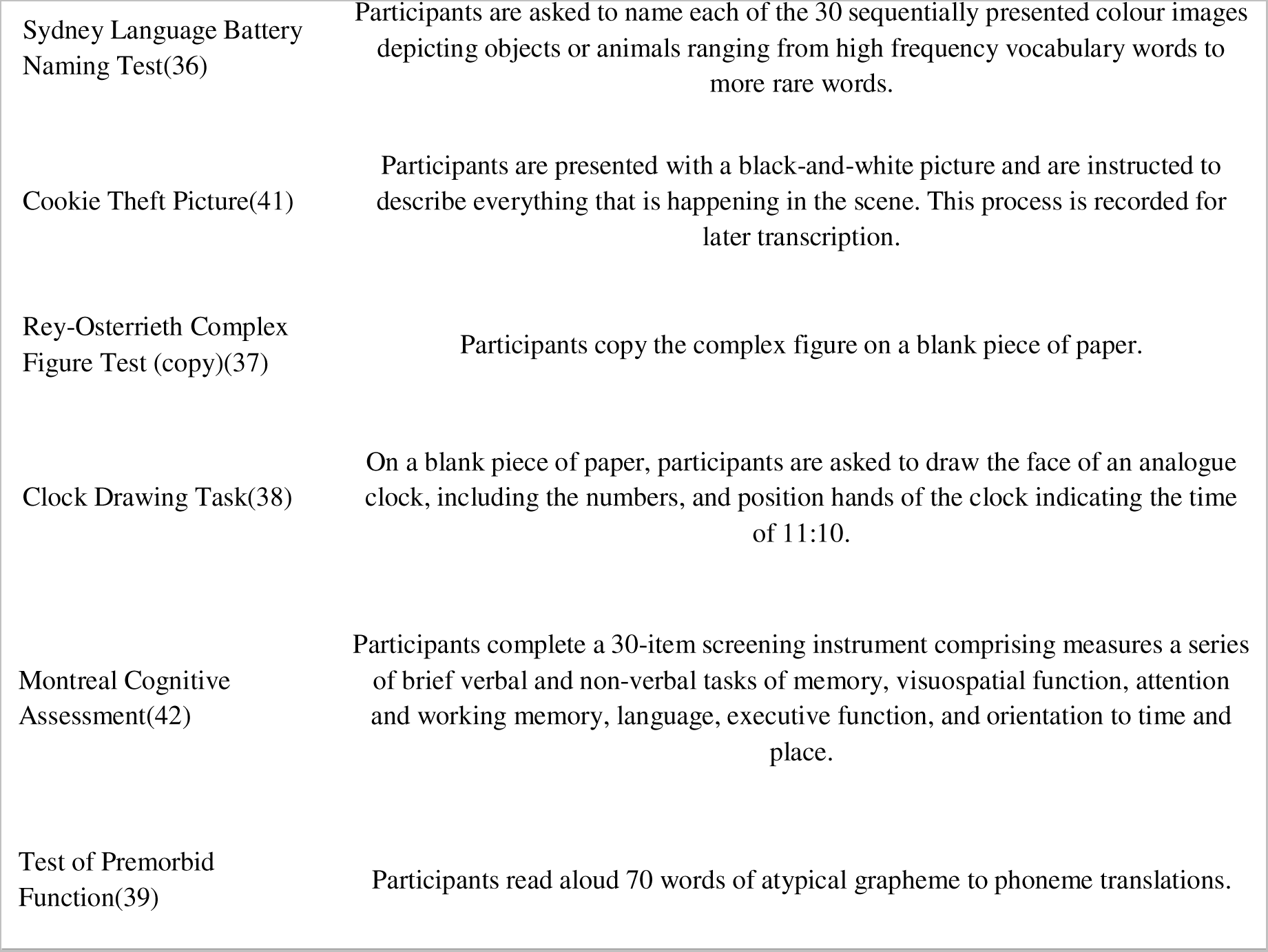
Description of cognitive instruments.

| Test name | Description |
| --- | --- |
| Digit Span (forward and backward)(32) | On the forward subtest, participants listen to strings of numbers of progressively increasing length and repeat the digits in the order of presentation. On the backward subtest, participants repeat numbers in reverse order. |
| Symbol Digit Modalities Test (oral version)(31) | The test comprises a coding key of nine abstract symbols, each paired with a one-digit number. The test sheet outlines the abstract symbols only, and participants verbally identify their corresponding numbers one-by-one by looking at the coding key. The goal is to identify as many numbers as possible in 90 seconds. |
| Trail Making Test(28) | In the part A, participants use a pen to connect 25 numbers in an ascending order as quickly as possible. In the part B, participants alternate between connecting 13 accenting numbers and 12 letters in the alphabetical order. |
| Victoria Stroop Test(30) | Participants are initially presented with 24 coloured dots and are asked to name colours of the dots as rapidly as possible. In the second part, they name ink colour of 24 neutral words. In the final part, participants name the colour of the ink for words with incongruent verbal content as quickly as possible. |
| Brief Visuospatial Memory Test-Revised(35) | Participants view a display with six geometric figures arranged in a 2 x 3 array for 10 seconds. Once the display is removed, they are asked to draw the figures in their correct location using paper-and-pencil. The test comprises a total of three learning trials. After an approximate 25-minute delay, participants draw the figures again from memory. On a subsequent recognition trial, participants are sequentially presented with 12 geometrical figures and identify which of the figures were on the original display. |
| Rey Auditory Verbal Learning Test(34) | Participants hear a list of 15 nouns and are asked to recall as many as they can at the end of each of the five learning trials. They are then read a new list of 15 nouns and are asked to recall as many as they can. Following the distractor list, participants recall the nouns from the first list. After a 20-minute delay, the first list is recalled again, followed by a recognition trial. |
| Verbal Fluency Test(33) | On the Letter Fluency subtest, participants generate words that begin with a specific letter (F, A, and S in the standard form; B, H, and R in the alternate form) as quickly as possible within one minute. Participants are instructed not to use names of people, places, or numbers, or to repeat the same word twice using a different ending. On the Category Fluency subtest, participants generate words that belong to a semantic category (animals and boys' names in the standard form; clothing and girls' names in the alternate form) as quickly as possible within one minute. |
| Sydney Language Battery<br>Naming Test(36) | Participants are asked to name each of the 30 sequentially presented colour images depicting objects or animals ranging from high frequency vocabulary words to more rare words. |
| Cookie Theft Picture(41) | Participants are presented with a black-and-white picture and are instructed to describe everything that is happening in the scene. This process is recorded for later transcription. |
| Rey-Osterrieth Complex<br>Figure Test (copy)(37) | Participants copy the complex figure on a blank piece of paper. |
| Clock Drawing Task(38) | On a blank piece of paper, participants are asked to draw the face of an analogue clock, including the numbers, and position hands of the clock indicating the time of 11:10. |
| Montreal Cognitive<br>Assessment(42) | Participants complete a 30-item screening instrument comprising measures a series of brief verbal and non-verbal tasks of memory, visuospatial function, attention and working memory, language, executive function, and orientation to time and place. |
| Test of Premorbid<br>Function(39) | Participants read aloud 70 words of atypical grapheme to phoneme translations. |

The raw scores are converted into standardized z-scores based on the demographically adjusted normative data. A test score was classified as impaired if the z-score <= −1.5 standard deviations.(27,40) A domain was classified as impaired if at least two scores were impaired (or one score for the visuospatial function domain) or a z-score averaged <= −1.5 across tests in that domain. A patient was classified as overall impaired if at least one domain was impaired.

#### Psycholinguistic analysis

Symptoms of speech and language dysfunction have commonly been documented acutely in ICANS. For example, Santomasso et al.(8) found that aphasia was observed as the first neurological feature in 19 of 22 patients with severe ICANS. Impaired naming, paraphasic errors, perseverative and hesitant speech were evident in 96% of these patients, and 19 of them rapidly transitioned to global aphasia. In the current study, all participants complete the Cookie Theft Picture Task (CTP),(41) which is audio recorded for transcription. The task is completed at baseline and at every follow-up, including acutely post-infusion. Psycholinguistic analysis of the recordings will identify linguistic markers specific to ICANS.

### Neurology assessment

All CAR-T patients complete standard care neurology assessments comprising clinical interview to collect medical history and elicit neurological complaint, physical neurological examination, and the Montreal Cognitive Assessment (MoCA)(42) as a screening measure of cognitive impairment. As part of standard of care, absolute values and patterns of laboratory markers (i.e., c-reactive protein, ferritin, lactic dehydrogenase, and white blood cell count including lymphocyte count) are collected pre- and post-CAR-T. The current project will review the available laboratory data to analyse its value in predicting risk of ICANS.

## Discussion

When a patient is referred for neuropsychological or neurological opinion in the context of a cognitive complaint or neurological dysfunction following CAR-T therapy, a clinician faces a challenge of establishing aetiology and predicting a recovery trajectory. This research will characterise a neurocognitive syndrome of ICANS as distinguishable from symptoms accompanying other treatment regiments. Our findings will facilitate early recognition of the syndrome and subsequently improve timely intervention and referral pathways. The research will develop evidence-based instruments for detecting and monitoring neurotoxicity. Through comprehensive baseline examinations and a longitudinal follow-up, the study will capture patient journey for up to a year following CAR-T therapy. Understanding the evolution of ICANS and its recovery will improve prognostication and counseling of patients and their families. Identifying new predictive markers for CAR-T related complications will enhance implementation of appropriate preventative strategies.

This research will capture a holistic patient experience, including their psychological status, quality of life, and the impact of common physical dysfunction, such as poor sleep, fatigue, and pain. The findings will improve referral pathways to optimise daily function in this cohort. Collection of demographically matched healthy control group will allow for a robust definition of psychometric impairment distinguishable from cognitive fluctuation in the healthy population. The study aims to improve the overall clinical experience of haematology patients pre- and post-CAR-T therapy.

## Data availability statement

Anonymised data, and the standardized proforma used to extract information on patient demographics and clinical characteristics, can be shared on reasonable request from qualified investigators.

## Author contribution

V.K., C.B.M, H.R., MA.A., M.D., T.K., S.J.H., and I.R. contributed to research design. Data is collected by V.K., H.R., H.O., C.S., S.vd.L., M.D., and S.R. V.K., C.B.M., T.K., H.O., H.R., C.S., F.D., and S.M.L. are involved in analyses and/or interpretation of the findings. All authors contributed to the final manuscript and approved it for submission.

## Acknowledgements

We acknowledge and thank CAR-T patients and their families for their participation. We also thank medical and administrative teams at Peter MacCallum Cancer Centre for their patient care and logistical contribution to this study. All authors substantially contributed to this work, with specific contributions outline in the author contribution section below.

## References

1. Siegler EL, Kenderian SS. Neurotoxicity and Cytokine Release Syndrome After Chimeric Antigen Receptor T Cell Therapy: Insights Into Mechanisms and Novel Therapies. Front Immunol [Internet]. 2020 [cited 2023 May 29];11. Available from: https://www.frontiersin.org/articles/10.3389/fimmu.2020.01973

2. Rice J, Nagle S, Randall J, Hinson HE. Chimeric Antigen Receptor T Cell-Related Neurotoxicity: Mechanisms, Clinical Presentation, and Approach to Treatment. Curr Treat Options Neurol [Internet]. 2019 Jul 20 [cited 2023 May 29];21(8):40. Available from: 10.1007/s11940-019-0580-3

3. Santomasso B, Bachier C, Westin J, Rezvani K, Shpall EJ. The Other Side of CAR T-Cell Therapy: Cytokine Release Syndrome, Neurologic Toxicity, and Financial Burden. Am Soc Clin Oncol Educ Book [Internet]. 2019 May [cited 2023 Jan 10];(39):433–44. Available from: https://ascopubs.org/doi/full/10.1200/EDBK_238691

4. Locke FL, Miklos DB, Jacobson CA, Perales MA, Kersten MJ, Oluwole OO, et al. Axicabtagene Ciloleucel as Second-Line Therapy for Large B-Cell Lymphoma. N Engl J Med [Internet]. 2022 Feb 17 [cited 2023 Jul 13];386(7):640–54. Available from: 10.1056/NEJMoa2116133

5. Tallantyre EC, Evans NA, Parry-Jones J, Morgan MPG, Jones CH, Ingram W. Neurological updates: neurological complications of CAR-T therapy. J Neurol [Internet]. 2021 Apr 1 [cited 2023 May 30];268(4):1544–54. Available from: 10.1007/s00415-020-10237-3

6. Grant SJ, Grimshaw AA, Silberstein J, Murdaugh D, Wildes TM, Rosko AE, et al. Clinical Presentation, Risk Factors, and Outcomes of Immune Effector Cell-Associated Neurotoxicity Syndrome Following Chimeric Antigen Receptor T Cell Therapy: A Systematic Review. Transplant Cell Ther [Internet]. 2022 Jun 1 [cited 2023 Jul 13];28(6):294–302. Available from: https://www.sciencedirect.com/science/article/pii/S2666636722001531

7. Holtzman NG, Xie H, Bentzen S, Kesari V, Bukhari A, El Chaer F, et al. Immune effector cell–associated neurotoxicity syndrome after chimeric antigen receptor T-cell therapy for lymphoma: predictive biomarkers and clinical outcomes. Neuro-Oncol [Internet]. 2021 Jan 1 [cited 2023 Jul 13];23(1):112–21. Available from: 10.1093/neuonc/noaa183

8. Santomasso BD, Park JH, Salloum D, Riviere I, Flynn J, Mead E, et al. Clinical and Biological Correlates of Neurotoxicity Associated with CAR T-cell Therapy in Patients with B-cell Acute Lymphoblastic Leukemia. Cancer Discov [Internet]. 2018 Aug 2 [cited 2023 Jul 13];8(8):958–71. Available from: 10.1158/2159-8290.CD-17-1319

9. Lee DW, Santomasso BD, Locke FL, Ghobadi A, Turtle CJ, Brudno JN, et al. ASTCT Consensus Grading for Cytokine Release Syndrome and Neurologic Toxicity Associated with Immune Effector Cells. Biol Blood Marrow Transplant [Internet]. 2019 Apr 1 [cited 2023 Jul 13];25(4):625–38. Available from: https://www.sciencedirect.com/science/article/pii/S1083879118316914

10. Herr MM, Chen GL, Ross M, Jacobson H, McKenzie R, Markel L, et al. Identification of Neurotoxicity after Chimeric Antigen Receptor (CAR) T Cell Infusion without Deterioration in the Immune Effector Cell-Associated Encephalopathy (ICE) Score. Biol Blood Marrow Transplant [Internet]. 2020 Nov 1 [cited 2023 May 29];26(11):e271–4. Available from: https://www.sciencedirect.com/science/article/pii/S1083879120304626

11. Schroyen G, Meylaers M, Deprez S, Blommaert J, Smeets A, Jacobs S, et al. Prevalence of leukoencephalopathy and its potential cognitive sequelae in cancer patients. J Chemother [Internet]. 2020 Oct 2 [cited 2023 May 29];32(7):327–43. Available from: 10.1080/1120009X.2020.1805239

12. Joly F, Castel H, Tron L, Lange M, Vardy J. Potential Effect of Immunotherapy Agents on Cognitive Function in Cancer Patients. JNCI J Natl Cancer Inst [Internet]. 2020 Feb 1 [cited 2023 May 29];112(2):123–7. Available from: 10.1093/jnci/djz168

13. Murrough JW, Iacoviello B, Neumeister A, Charney DS, Iosifescu DV. Cognitive dysfunction in depression: Neurocircuitry and new therapeutic strategies. Neurobiol Learn Mem [Internet]. 2011 Nov 1 [cited 2023 May 29];96(4):553–63. Available from: https://www.sciencedirect.com/science/article/pii/S1074742711001171

14. Murri MB, Caruso R, Christensen AP, Folesani F, Nanni MG, Grassi L. The facets of psychopathology in patients with cancer: Cross-sectional and longitudinal network analyses. J Psychosom Res [Internet]. 2023 Feb 1 [cited 2023 May 29];165:111139. Available from: https://www.sciencedirect.com/science/article/pii/S002239992200424X

15. Clinton-McHarg T, Carey M, Sanson-Fisher R, Tzelepis F, Bryant J, Williamson A. Anxiety and depression among haematological cancer patients attending treatment centres: Prevalence and predictors. J Affect Disord [Internet]. 2014 Aug 20 [cited 2023 May 29];165:176–81. Available from: https://www.sciencedirect.com/science/article/pii/S0165032714002663

16. Rivera AM, May S, Lei M, Qualls S, Bushey K, Rubin DB, et al. CAR T-Cell-Associated Neurotoxicity: Current Management and Emerging Treatment Strategies. Crit Care Nurs Q [Internet]. 2020 Apr [cited 2023 Jul 22];43(2):191–204. Available from: https://journals.lww.com/10.1097/CNQ.0000000000000302

17. Maillet D, Belin C, Moroni C, Cuzzubbo S, Ursu R, Sirven-Villaros L, et al. Evaluation of mid-term (6-12 months) neurotoxicity in B-cell lymphoma patients treated with CAR T cells: a prospective cohort study. Neuro-Oncol [Internet]. 2021 Sep 1 [cited 2023 Jan 28];23(9):1569–75. Available from: 10.1093/neuonc/noab077

18. Wang XS, Srour SA, Whisenant M, Subbiah IM, Chen TH, Ponce D, et al. Patient-Reported Symptom and Functioning Status during the First 12 Months after Chimeric Antigen Receptor T Cell Therapy for Hematologic Malignancies. Transplant Cell Ther [Internet]. 2021 Nov 1 [cited 2023 Jul 22];27(11):930.e1–930.e10. Available from: https://www.sciencedirect.com/science/article/pii/S2666636721010691

19. Gust J, Hay KA, Hanafi LA, Li D, Myerson D, Gonzalez-Cuyar LF, et al. Endothelial Activation and Blood–Brain Barrier Disruption in Neurotoxicity after Adoptive Immunotherapy with CD19 CAR-T Cells. Cancer Discov [Internet]. 2017 Dec 4 [cited 2023 Aug 21];7(12):1404–19. Available from: 10.1158/2159-8290.CD-17-0698

20. Greenbaum U, Strati P, Saliba RM, Torres J, Rondon G, Nieto Y, et al. CRP and ferritin in addition to the EASIX score predict CAR-T–related toxicity. Blood Adv [Internet]. 2021 Jul 15 [cited 2023 Aug 21];5(14):2799–806. Available from: 10.1182/bloodadvances.2021004575

21. Amidi Y, Eckhardt CA, Quadri SA, Malik P, Firme MS, Jones DK, et al. Forecasting immune effector cell-associated neurotoxicity syndrome after chimeric antigen receptor t-cell therapy. J Immunother Cancer [Internet]. 2022 Nov 1 [cited 2023 Aug 21];10(11):e005459. Available from: https://jitc.bmj.com/content/10/11/e005459

22. Blais MA, Sinclair SJ. Introduction to the SPECTRA: Indices of Psychopathology:

23. Blais MA, Sinclair SJ, Richardson LA, Massey C, Stein MB. External correlates of the SPECTRA: Indices of psychopathology (SPECTRA) in a clinical sample. Clin Psychol Psychother [Internet]. 2021 [cited 2023 May 30];28(4):929–38. Available from: https://onlinelibrary.wiley.com/doi/abs/10.1002/cpp.2546

24. Cella D, Riley W, Stone A, Rothrock N, Reeve B, Yount S, et al. The Patient-Reported Outcomes Measurement Information System (PROMIS) developed and tested its first wave of adult self-reported health outcome item banks: 2005–2008. J Clin Epidemiol [Internet]. 2010 Nov 1 [cited 2023 Jul 25];63(11):1179–94. Available from: https://www.sciencedirect.com/science/article/pii/S0895435610001733

25. Herdman M, Gudex C, Lloyd A, Janssen MF, Kind P, Parkin D, et al. Development and preliminary testing of the new five-level version of EQ-5D (EQ-5D-5L). Qual Life Res [Internet]. 2011 Dec 1 [cited 2023 Jul 25];20(10):1727–36. Available from: 10.1007/s11136-011-9903-x

26. Buckley RF, Ellis KA, Ames D, Rowe CC, Lautenschlager NT, Maruff P, et al. Phenomenological characterization of memory complaints in preclinical and prodromal Alzheimer’s disease. Neuropsychology. 2015;29:571–81.

27. Wefel JS, Vardy J, Ahles T, Schagen SB. International Cognition and Cancer Task Force recommendations to harmonise studies of cognitive function in patients with cancer. Lancet Oncol [Internet]. 2011 Jul 1 [cited 2023 May 29];12(7):703–8. Available from: https://www.sciencedirect.com/science/article/pii/S1470204510702941

28. Reitan RM. The Relation of the Trail Making Test to Organic Brain Damage. Journal of Consulting Psychology. 1955;19(5):393–4.

29. Tombaugh TN. Trail Making Test A and B: Normative data stratified by age and education. Arch Clin Neuropsychol [Internet]. 2004 Mar 1 [cited 2023 May 30];19(2):203–14. Available from: https://www.sciencedirect.com/science/article/pii/S0887617703000398

30. Regard M. Cognitive rigidity and flexibility: A neuropsychological study. Victoria: University of Victoria; 1981.

31. Smith A. Symbol digit modalities test. Los Angeles: Western psychological services; 1973. 22 p.

32. Wechsler D. Wechsler Adult Intelligence Scale: Technical and interpretive manual. 4th ed. San Antonio: Pearson Assessment; 2008.

33. Delis D, Kaplan E, Kramer J. Delis-Kaplan executive function system. Assessment; 2001.

34. Lezak M. Neuropsychological assessment. 2nd ed. New York: Oxford University Press; 1983.

35. Benedict R. Brief visuospatial memory test--revised. PAR; 1997.

36. Savage S, Hsieh S, Leslie F, Foxe D, Piguet O, Hodges JR. Distinguishing subtypes in primary progressive aphasia: application of the Sydney language battery. Dement Geriatr Cogn Disord [Internet]. 2013 [cited 2023 May 30];35(3–4):208–18. Available from: https://search.ebscohost.com/login.aspx?direct=true&AuthType=sso&db=mnh&AN=234673 07&site=ehost-live&custid=s2775460

37. Meyers JE, Meyers KR. Rey complex figure test under four different administration procedures. Clin Neuropsychol [Internet]. 1995 Feb 1 [cited 2023 May 30];9(1):63–7. Available from: 10.1080/13854049508402059

38. Shulman KI, Shedletsky R, Silver IL. The challenge of time: Clock-drawing and cognitive function in the elderly. Int J Geriatr Psychiatry [Internet]. 1986 [cited 2023 Jul 25];1(2):135–40. Available from: https://onlinelibrary.wiley.com/doi/abs/10.1002/gps.930010209

39. Wechsler D. Advanced clinical solutions for the WAIS-IV and WMS-IV. San Antonio: The Psychological Corporation; 2009.

40. Jak AJ, Bondi MW, Delano-Wood L, Wierenga C, Corey-Bloom J, Salmon DP, et al. Quantification of Five Neuropsychological Approaches to Defining Mild Cognitive Impairment. Am J Geriatr Psychiatry [Internet]. 2009 May 1 [cited 2023 Jan 11];17(5):368–75. Available from: https://www.sciencedirect.com/science/article/pii/S106474811260743X

41. Goodglass H, Kaplan E, Weintraub S. BDAE: The Boston diagnostic aphasia examination. Philadelphia, PA: Lippincott Williams & Wilkins; 2001.

42. Nasreddine ZS, Phillips NA, Bédirian V, Charbonneau S, Whitehead V, Collin I, et al. The Montreal Cognitive Assessment, MoCA: A Brief Screening Tool For Mild Cognitive Impairment. J Am Geriatr Soc [Internet]. 2005 [cited 2023 Mar 31];53(4):695–9. Available from: https://onlinelibrary.wiley.com/doi/abs/10.1111/j.1532-5415.2005.53221.x

